# ‘Truthsets’ for clinical validation of large-scale functional assays: Practice recommendations from Cancer Variant Interpretation Group UK (CanVIG-UK)

**DOI:** 10.64898/2026.07.10.26357770

**Authors:** Sophie Allen, Charlie F. Rowlands, Alice Garrett, Zeid Kuzbari, Miranda Durkie, George J. Burghel, Rachel Robinson, Alison Callaway, Joanne Field, Bethan Frugtniet, Sheila Palmer-Smith, Jonathan Grant, Judith Pagan, Elizabeth Johnston, Trudi McDevitt, Lowri Hughes, Laura Yarram-Smith, Peter Logan, Laura Reed, Katie Snape, Terri McVeigh, Helen Hanson, Rehan Villani, Amanda B. Spurdle, Lea M. Starita, Douglas M Fowler, Frederick P. Roth, Elizabeth Radford, David J. Adams, Gregory M. Findlay, Clare Turnbull, CanVIG-UK

## Abstract

**Background:** Large-scale functional assays, including multiplex assays of variant effect, have substantial potential to resolve variants of uncertain significance (VUS), particularly for rare missense variants where clinical and population evidence are limited. The ClinGen assay-level clinical validation framework described by Brnich et al provided baseline guidance for the use of functional data for variant classification. However, clear consensus regarding construction of variant ‘truthsets’ by which to clinically validate functional data remains lacking.

**Methods:** CanVIG-UK developed consensus recommendations for truthset construction through an iterative national consultation process involving the CanVIG Steering Advisory Group (CStAG), wider CanVIG-UK membership, and engagement with international functional genomics experts. Consultation was based on previous analyses of 2,120 truthset constructions examining the impact of truthset composition on evidence point allocation within the ClinGen assay-level clinical validation framework.

**Results:** Across several consultations, CanVIG-UK established nine guiding principles and seven best-practice recommendations for assay-level clinical validation, using the assumed context of an assay for a cancer susceptibility gene where loss-of-function is the mechanism of pathogenicity. The principal recommendation stipulates, where assays are intended for use in interpretation of largely missense variants, the truthset used to validate should comprise only missense variants. Rather than mixtures of different variant types which may serve to over-estimate assay performance. Additional recommendations support option for relaxation of truthset stringency to improve power, augmentation of benign missense truthsets with systematically derived ‘proxy-clinical’ benign variants, independent clinical validation separate from assayist-defined validation, and careful evaluation of missense score distributions against that of protein-truncating and synonymous variants. Guidance is also provided for scenarios with limited pathogenic truthset availability and for assays reporting multiple deleterious zones or readouts.

**Conclusions:** The CanVIG-UK principles and recommendations for truthset construction upon the ClinGen assay-level clinical validation framework, while aiming to form a baseline for future discussion regarding other functional and disease contexts and helping to address the gap between publication of new data and routine clinical implementation.

## Background

The continuing rise in the quantity of variants classified as being of uncertain significance (VUS) is well documented(1). Despite larger volumes of population sequencing data demonstrating more variants as being recurrently present in the normal population, the volume of rare variants for which there are insufficient clinical data by which to classify the variant also continues to increase, disproportionately so for individuals of non-European ancestry for whom available clinical and population data are particularly sparse.

Robust functional data offer rich opportunity for classification of VUS, since results are not reliant on clinical observations or population data, and can be produced for any number of previously unseen variants. The availability of large-scale multiplex assays of variant effect is increasing at a rapid rate, providing transformative opportunity for resolution of VUS, particularly for variants arising in non-European populations(2, 3).

Guidance for application to variant classification of functional assay data was included in the American College of Medical Genetics and Association for Molecular Pathology (ACMG/AMP) 2015 recommendations (hereafter termed v3.0), recommending application of ‘strong’ evidence towards pathogenicity (PS3) and benignity (BS3) for “well-established” assays (4). In the UK, these guidelines were in 2016 adopted and adapted into the Association for Clinical Genomic Science (ACGS) Best Practice Guidelines for Variant Classification in Rare Disease(5), and in 2017 the Cancer Variant Interpretation Group UK (CanVIG-UK) was initiated, a multidisciplinary group with the primary goal of driving consistent implementation within UK NHS diagnostic science of the ACMG/AMP v3.0 guidelines for variant classification in cancer susceptibility genes (6, 7).

Following the v3.0 recommendations, subsequent papers by Tavtigian et al. in 2018 and 2020 proved that the qualitative evidence strengths described in the v3.0 guidelines (such as ‘strong’) are compatible with a naïve-Bayesian classifier, thus assigning each evidence strength quantitative ‘evidence points’ (hereafter EPs), where ‘strong’ is equivalent to 4 EPs allocatable towards pathogenicity or benignity(8, 9).

The 2020 ClinGen publication by Brnich et al. expanded on this guidance for functional data, providing a framework to guide clinical review of functional assays and introducing use of clinical validation controls, whereby the evidence strength applicable for an assay depends on the assay correctly identifying reference sets of clinically-classified pathogenic variants (a ‘pathogenicity truthset’) as deleterious and reference sets of clinically-classified benign variants (a ‘benignity truthset’) as neutral. Brnich et al. highlighted need for pre-definition of a threshold delineating deleterious from neutral, and that this typically would be as supplied by the assayist from initial calibration of the assay. In the most typical context of an assay quantifying loss-of-function, the deleterious assay readouts are typically termed non-functional and the neutral readouts termed functional(10). Assay concordance could then be assessed by calculating a quantitative posterior probability that could be converted to EPs following the same conversion as published by Tavtigian *et al*. Notably, the direction of EPs (PS3 versus BS3) is conversely related to the truthset under examination. So EPs for pathogenicity (PS3) are primarily driven by concordance of clinically-classified benign variants with neutral (functional) assay readouts; EPs for benignity (BS3) are primarily driven by concordance of clinically-classified pathogenic variants with deleterious (non-functional) assay readouts(11).

Using this system, evidence scoring fits onto a continuous scale, an upgrade from the categorical evidence system described in the original 2015 ACMG/AMP v3.0 framework of supporting/moderate/strong/very strong. Furthermore, in the upcoming v4.0 release of the ACMG/AMP/CAPP/ClinGen guidelines, functional evidence will be applicable on a scale from -8 to +8 points. Given that in the v4.0 framework, an overall score of -1 (for Likely Benign) or +6 (for Likely Pathogenic) will be sufficient for classification out of VUS regardless of the provenance of the evidence points, this means there will be potential for classification to become feasible just on the basis of functional data.

The EPs allocatable from an assay are inherently determined by the methodology used for clinical validation of the assay; a key element in clinical validation is delineation of what variants are appropriate to constitute truthsets. In the 2020 ClinGen framework from Brnich *et al*, they state broadly that “controls should also be relevant to the disease mechanism (such as gain-of-function or loss-of-function) and the type of variant under consideration (e.g., missense controls for evaluating missense variants of uncertain significance)”(2). Whilst missense variants are the type for which functional evidence is clearly most likely to influence the classification, use of missense variants for clinical validation is not explicitly mandated by Brnich et al, leaving ambiguity and discordance regarding this issue. Where assayists have conducted a clinical validation as part of their assay publication, there is wide variety in the composition of pathogenic and benign truthsets presented, which may in part reflect motivation to optimise presentational validation performance and therefore perceived clinical utility of their assay.

In both the UK and internationally, this has led to a lack of confidence amongst clinical diagnostic scientists and laboratories regarding assayist-published assay validations(12–15). Mixed with a lack of confidence in conducting such clinical validations themselves, a problematic hiatus has emerged between publication of new functional data and assimilation of the data by clinical diagnostic services for clinical variant classification. There is therefore urgent requirement within our community for clear rules that will enable independent, consistent truthset construction and clinical validation to address this implementation gap.

Hence, CanVIG-UK sought to build upon the Brnich *et al.,* 2020 ClinGen assay-level clinical validation framework to further define best practice for truthset construction and application in clinical validation of functional assays. We first conducted a survey of current practices in truthset assembly, examining CSpec pages for 112 genes across 39 VCEPs, elucidating how (i) functional data are inconsistently applied and (ii) how the provenance of and rules for establishment of truthsets vary widely (hereafter termed the CSpec survey). Next, examining four genes and corresponding assays, we performed 2,120 different clinical validations to explore how differing sets of rules for truthset construction impacted the size of truthset assembled and the consequent EPs (hereafter termed the truthset analyses)(16). These results are published separately but are summarised in Box 1. Then, through a series of national CanVIG-UK meetings and intervening meetings of the CanVIG Steering Advisory Group (CStAG), we iteratively evolved a series of consensus principles and best practice recommendations by which to govern CanVIG-UK standards for truthset construction and application in clinical validation of functional assays

## Methods

The Cancer Variant Interpretation Group UK (CanVIG-UK) comprises >450 members, with 75-150 attendees for each monthly online meeting (duration 1.25 hours). The CanVIG Steering Advisory Group (CStAG) has oversight of CanVIG-UK, comprises 18 senior clinical scientists and consultant clinical geneticists in total representing all England Genomic Laboratory Hubs, each devolved nations (Scotland, Wales, Northern Ireland), and the Republic of Ireland and meets separately each month (1.75 hours).

The preliminary findings from the truthset analysis were presented to CStAG on 26^th^ June 2025 and to CanVIG-UK on 10^th^ October 2025, along with an instructional session on the ClinGen assay-level clinical validation framework and paradoxical impact of truthset size. Evolution of the truthset analyses, principles and recommendations was based on these discussions. Updated analyses, principles and recommendations were then presented to CanVIG-UK 13th February 2026, and to CStAG on 26^th^ February 2026, 26th March 2026, and 28th May 2026. Repeated iterations of the truthset analyses and recommendations were reviewed, with draft versions of the final recommendations circulated and ratified within CStAG and CanVIG-UK (17^th^ February 2026, 15^th^ June 2026).

The preliminary findings from the truthset analysis were also presented to the ClinGen Functional working Group on 16th September 2025 and the recommendations on 16^th^ June 2026; the truthset analyses and recommendations were multiply reviewed by AS, RV, LS, FR, ER and DF on 20^th^ February 2026, 25^th^ March 2026 and 16^th^ June 2026. Although not members of CanVIG-UK or CStAG, review and evaluation by the ClinGen FWG and AVE Alliance expert international stakeholders was sought to ensure we were maintaining consistency between the consensus CanVIG-UK recommendations and the emerging recommendations from the ClinGen FWG.

#### Box 1: Observations from analyses of 2,120 gene-truthset-assay dyads. Full details of this analysis are published separately(16)

Instructive to development by CanVIG-UK of its principles and recommendations were findings from analyses of 2120 gene-assay-truthset combinations (available in full detail in Allen et al., 2026(16)). In summary, the impact on allocatable EPs using the ClinGen assay-level clinical validation framework described by Brnich et al was compared when adjusting truthset composition by various parameters. The findings overall reflected how predictiveness (concordance) and confidence (power, namely number of available truthset variants) will vary with different truthset compositions. The following parameters for truthset composition were assessed:

1. **Variant type** (Missense vs PTV/Synonymous vs All variants). Missense variants typically resulted in lower allocatable EPs, largely reflecting lower power along with modestly lower concordance (see Figure 1).
2. **Stringency** regarding (i) ClinVar classification strength (e.g. likely pathogenic vs pathogenic), (ii) ClinVar submitters (no classification criteria (0*) vs single submitter (1*) vs multiple submitters (2*) vs expert panel submission (3*)), (iii) Allowance of ‘soft-conflicts’ (i.e. a conflicting classification between between ClinVar submitters, for example, likely pathogenic and VUS). Higher stringency for any of the measures typically resulted in reduced allocatable EPs due to reduction in power that was of greater impact than any increased concordance.
3. **Augmentation of ClinVar-classified benign missense truthsets** with ‘proxy-clinical’ benign-classified missense variants, which have not been clinically reported but have been systematically assembled by applying ACMG/AMP rules to available data sources (eg population data from gnomAD v4.1 and in silico predictions). Inclusion of ‘proxy-clinical’ benign-classified missense variants resulted in increased allocatable EPs for the majority of gene-assay dyads. Varying the stringency of the rules for proxy-clinical benignity classification caused different trade-offs of concordance and power for different gene-assay dyads (Supplementary Table 1)
4. **Reported phenotypic association** for ClinVar-classified pathogenic variants. Using a tighter phenotypic description typically resulted in lower allocatable EPs; again this was typically due to lower power despite modestly improved concordance
5. **Deleterious score zone for assays with multiple reported deleterious zones**. Clinical validation for the most deleterious zone only, assigning the weaker—scoring deleterious zone as intermediate, typically optimised the allocatable EPs towards both pathogenicity and benignity (albeit creating a wider non-scoring intermediate zone).

**Figure 1:**
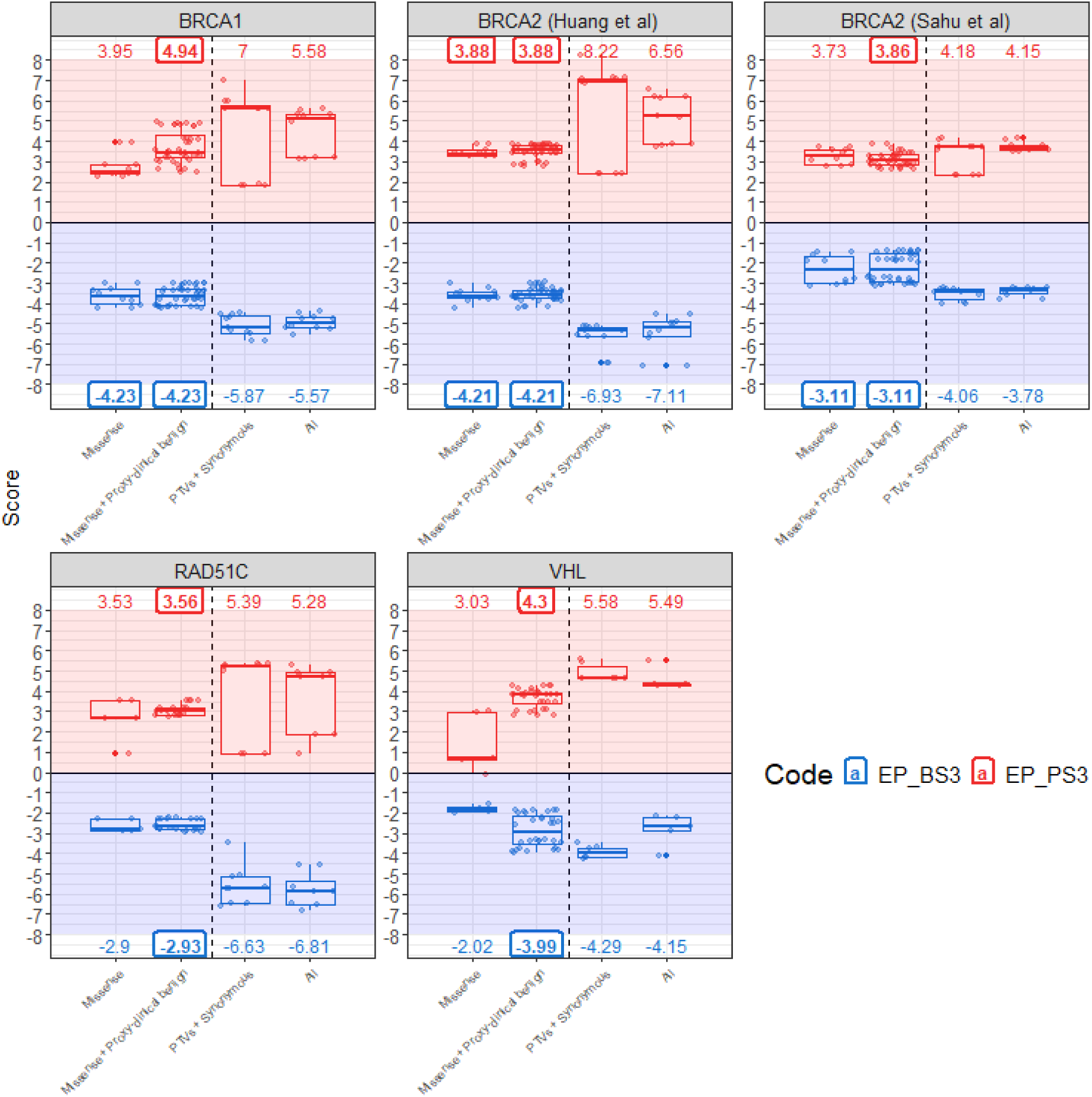
Maximum allocatable EPs towards pathogenicity and benignity from comparison of 70 approaches to ClinVar-based truthset assembly for each of five assays. For each gene, four high-level approaches to ClinVar-extracted truthset types are presented. The range of allocatable EPs using all examined missense-only truthsets (either ClinVar-extracted only, or ClinVar plus proxy-clinical benign-classified missense variants) are illustrated in the first two bars for each assay plot, with results for PTV/synonymous and All ClinVar variants presented for comparison. Maximum scores are highlighted in boxes above the relevant assay type, and further details about the provenance of the highest scoring missense truthset are provided in Supplementary Table 1. PTV=Protein-Truncating Variant, EP_BS3=EPs for benignity, EP_PS3=EPs for pathogenicity. Impact of truthset composition on EPs was explored for five assays (BRCA1=Findlay et al., 2018; BRCA2_Huang=Huang et al., 2025; BRCA2_Sahu=Sahu et al., 2025; RAD51C=Olvera-León et al., 2024; VHL=Buckley et al., 2024(17–21).

## Results

Via iterative consultation across CStAG and CanVIG-UK and the expert international stakeholders, and based on the findings from the truthset analysis and CSpec survey, we established nine principles relating to truthset assembly for assay-level clinical validation using the ClinGen assay-level clinical validation framework described by Brnich et al., 2020(10). These principles informed our subsequently presented Best Practice Recommendations for assay-level clinical validation. CanVIG-UK guidance represents recommended practice for approaches considered appropriate for routine clinical implementation. The term **aspirational best practice** is used for approaches considered optimal, but which may not currently be feasible or achievable due to variant numbers and/or data quality. Whilst broadly applicable to all genes and assay types, these principles and recommendations were developed considering assays for cancer susceptibility genes acting via loss-of-function. Some specific factors relating to other assay types are listed at the end as ancillary recommendations

### Principles for truthset assembly for assay clinical validation

#### Principle A: Quantitative clinical validation of a functional assay should reflect not just the predictiveness of the assay against reference controls (truthsets) but also the confidence of that predictiveness

Predictiveness is reflected by the concordance of assay readouts with truthset classifications. Confidence is determined by the number of truthset variants with which the evaluation of concordance was undertaken. Within the ClinGen assay-level clinical validation framework described by Brnich et al, incorporation of a ‘+1’ correction to the posterior probabilities embeds this reflection of power (confidence) in the EPs. Direct estimates of sensitivity and specificity measure predictiveness but without a reflection of our confidence therein.

#### Principle B: Variant assessment should reflect mechanistic asymmetry

Only one mechanism of action need be demonstrated as abrogated to qualify a variant as deleterious with subsequent classification of pathogenicity; “proving benignity” requires that all possible mechanisms must be examined to confirm a variant is neutral. An assay may capture only a single or subset of the mechanisms by which a variant can be deleterious to protein function. In theory, there will always be an aspect of biological function not recapitulated by an assay, be it mechanism, cell type or developmental stage. Thus, there inherently is some element of doubt to a neutral (functional) assay readout.

#### Principle C: Classification of variants from VUS to pathogenic is of greater clinical utility; but also where errors are likely to be most consequential

For a “cold VUS”, that is a variant with minimal additional attached evidence, if PS3 from functional data enables a clinically actionable classification shift to (likely) pathogenic, conferment of a genetic diagnosis will typically result in alteration to management and familial cascading to assign family members as at-risk variant carriers. The corollary of which is that the harm is likely significantly substantial if the functional evidence transpires to be erroneous and if the classification then requires down-grading. If the “cold VUS” is classified as (likely) benign through conferment of BS3, no new clinical actions ensue meaning adverse consequences are likely limited if it transpires this was erroneous.

#### Principle D: Missense variants are the variant type for which functional data are most likely to influence classification

Because it is of high *a priori* likelihood that a PTV will be pathogenic and that a synonymous variant will be benign, little additional evidence is required for these variant types to achieve these classifications. Conversely, rare missense variants are much more likely to reside as VUS, as they typically lack the substantive clinical evidence required to attain classification as pathogenic; classification of a rare missense variant as benign is also challenging under ACMG v3.0 rules as two evidence types are required.

Accordingly, missense is the variant type for which EPs from assay data are most likely to influence clinically actionable classification. Thus, whilst it is a valuable first step to demonstrate that the assay can distinguish complete loss of function (PTVs) from normal function (synonymous variants), the critical question is “can the assay distinguish pathogenic missense variants from benign missense variants?” A MAVE could perfectly separate PTVs from synonymous variants and still perform poorly on missense variants if (i) assay readouts for pathogenic missense variants are of more modest deleteriousness, (ii) benign missense variants produce mild functional perturbations on assay and/or (iii) the assay captures one aspect of protein function but not others.

Pathogenic and benign missense controls thus can be argued to provide the most appropriate validation framework because they evaluate assay performance on the same variant class and clinical decision space for which evidence will ultimately be applied. Protein-truncating and synonymous variants remain valuable mechanistic controls for establishing assay responsiveness to loss-of-function and normal-function states, respectively, but may overestimate assay performance if used as the primary truthset for clinical validation where applied primarily for interpretation of missense variants.

Furthermore, for assays not recapitulating genomic context, splicing effects and nonsense mediated decay are not captured, meaning assay readouts for nonsense and splicing PTVs may be neutral.

#### Principle E: Assayist-defined thresholds for deleterious and neutral will impact downstream clinical validation

Aspirational best practice would be that assay calibration and validation were performed using two independent but equivalent truthsets; for example, two independent sets of clinically-classified missense variants each comprising both benign and pathogenic classifications.

However, current operational application of the ClinGen framework generally relies upon leveraging of the assayist-defined thresholds. Typically these are derived using sets of PTVs and synonymous variants, applying maximum likelihood estimation or false discovery rates to define the best-fitting threshold between neutral and deleterious, yielding as parsimonious as possible an ‘intermediate’ zone.

For assays for which separation in the calibration was very clean, there may be a substantive ‘bald’ zone in between the clusters of positive and negative controls (typically PTVs and synonymous variants), with, if any, a very narrow intermediate zone. The distribution of pathogenic and benign missense variants in relation to the thresholds may not directly match that of calibration positive and negative controls (Box 2).

#### Principle F: The lack of clinically-classified benign missense variants is problematic

This paucity is problematic because the number of available benign missense variants will mostly influence EPs allocatable for pathogenicity. As per Principle C, assay EPs allocatable for pathogenicity will have the highest impact on clinically actionable classification shifts. This paucity likely reflects laboratories (i) not routinely wrangling between classification as cold VUS versus likely benign (especially if neither such variants are included on clinical reports), and (ii) not depositing into ClinVar clinical classifications for (likely) benign variants. High stringency benignity truthset classifications are particularly sparse, in particular ClinVar 2* classifications or 1* classifications free of soft-conflicts.

#### Principle G: The lack of clinically-classified pathogenic missense variants can also be problematic

For many more established genes this may be a less substantial issue numerically than for benign missense truthset variants, and BS3 allocation and classification as benignity is less likely to be clinically actionable. Nevertheless, this is definitely a limiting factor in particular for less frequently tested genes. Availability of robust clinically-classified pathogenic missense variant controls for validation is key to demonstrating with confidence that a call of neutral (functional) by the assay has excluded these mechanisms of action.

#### Principle H: Contamination of truthsets will attenuate EPs allocatable for the assay

Any contamination of truthsets with incorrect classification or hypomorphic variants will attenuate the allocatable EPs from clinical validation. This is due to regression towards the null. This will therefore only serve to *deflate* (under-score) the EPs compared to that which would be achieved from clinical validation using a hypothetical ‘perfect truthset’. Accordingly, if power dictates benefit in EPs from using lower stringency truthsets, this will not result in over-scoring of assays and/or erroneous over-allocation of EPs.

#### Principle I: There is potential for circularity in truthset construction where clinical variant classifications have relied on functional data

ClinVar classifications of pathogenicity or benignity may have been reached via the contribution of functional data. This creates circularity if the assay data used for classification is non-orthogonal with the assay under evaluation. Clearly, this is particularly significant where clinical validation is undertaken subsequent to publication and the ClinVar classifications available for truthsets may include functional data from the assay under evaluation.

### CanVIG-UK Best Practice Recommendations for assay-level clinical validation of functional assays

#### Recommendation 1: Truthsets used in clinical validation should comprise missense variants only

This applies to assays for which the readouts would primarily influencing classification of missense variants: the likely majority of scenarios.

Applying a truthset comprising ‘all ClinVar variants’ (i.e. an arbitrary mixture of missense/protein-truncating/synonymous variants) is not recommended. If it is recognised that performance of the assay against missense variants must be demonstrated, it is then illogical to propose clinical validation can deploy truthsets of missense mixed in with PTVs/synonymous variants where signal from missense discordance may be diluted out. This practice would thus risk over-estimation of assay predictiveness for missense variants where assay predictiveness for protein-truncating/synonymous variants did not translate to missense variants.

#### Recommendation 2: Clinical validation should be performed independently to initial validation undertaken by the original assayists

Aspirational best practice would be to ensure the clinical validation uses truthset variants distinct to those used as internal controls for assayist-defined thresholds calibration; this may not be feasible.

#### Recommendation 3: Stringency of truthsets can be relaxed thus improving power for increased allocatable Eps

The EPs for pathogenicity and benignity can be derived using truthsets of differing stringency/provenance, where this provides greater allocatable EPs (Figure 1, Supplementary Table 1). Truthsets should be constructed using prescribed criteria ahead of and agnostic to their performance in the assay (ie, the truthset should not be amended post-hoc to improve concordance).

#### Recommendation 4: Clinically-classified benign missense truthsets can be augmented with proxy-clinical benign-classified missense variants

These are missense variants which have not been clinically submitted to ClinVar as benign, but for which systematic ACMG/AMP v3.0 classification using population and in-silico data results in classification as (likely) benign(11). *Note:* Where there are no concordant pathogenic missense truthset variants, a mathematical fix in manner of a Haldane correction should be applied to enable the ClinGen assay-level clinical validation framework to enable calculation of EPs for pathogenicity based on the available benign missense truthset variants (see Table 1)

**Table 1:** Mathematical fix to enable application of pathogenic evidence using the ClinGen assay-level clinical validation framework from Brnich et al. As per Recommendation 4, where a) there are zero concordant variants in the pathogenic truthset, and b) there are sufficient benign variants in the truthset to confer allocatable EPs for pathogenicity (PS3). In this scenario (row 1), there are 0 pathogenic truthset variants and 2147 benign truthset variants, 1287 of which are concordant. *Insertion of a single pathogenic concordant ‘pseudo-truthset’ variant allows the validation framework calculation to yield the 4.33 EPs for pathogenicity using the concordance of the benign truthset.* Note that the number of concordant pseudo-truthset variants and size of the prior does not actionably impact the allocatable EPs for pathogenicity (rows 2-8). Clearly, in the scenario of applying this mathematical fix due to there being no concordant pathogenic truthset variants, *no EPs for benignity (BS3) should be allocatable.* TP=True Positive; FP=False Positive; FN=False Negative; TN=True Negative.

| Row | # Classified (truth) |  |  | Proportion pathogenic (Prior, P<sub>1</sub>) | Assay result |  |  |  |  |  | Plus 1 |  |  |  |  |  |
| --- | --- | --- | --- | --- | --- | --- | --- | --- | --- | --- | --- | --- | --- | --- | --- | --- |
|  | Total | Pathogenic | Benign |  | Deleterious |  | Intermediate |  | Neutral |  | "+1 proportions" (Posterior, P<sub>2</sub>) |  | OddsPath= [P<sub>2</sub> x (1- P<sub>1</sub>)] / [(1- P<sub>2</sub>) x P<sub>1</sub>] |  | EPs EP=log<sub>2.08</sub> OddsPath |  |
|  |  |  |  |  | Path (TP) | Benign (FP) | Path | Benign | Path (FN) | Benign (TN) | Pathogenic | Benign | Pathogenicity (PS3) | Benignity (BS3) | EP-PS3 | EP-BS3 |
| 1 | 2147 | 0 | 2147 | 0 | 0 | 89 | 0 | 771 | 0 | 1287 | 0.00 | 1.00 | NA | NA | NA | NA |
| 2 | 2148 | 1 | 2147 | 0.00047 | 1 | 89 | 0 | 771 | 0 | 1287 | 0.01 | 1.00 | 23.86 | 0.60 | 4.331 | 0.699 |
| 3 | 2149 | 2 | 2147 | 0.00093 | 2 | 89 | 0 | 771 | 0 | 1287 | 0.02 | 1.00 | 23.86 | 1.20 | 4.331 | 0.248 |
| 4 | 2152 | 5 | 2147 | 0.00232 | 5 | 89 | 0 | 771 | 0 | 1287 | 0.05 | 1.00 | 23.86 | 1.80 | 4.331 | 0.801 |
| 5 | 2157 | 10 | 2147 | 0.00464 | 10 | 89 | 0 | 771 | 0 | 1287 | 0.10 | 1.00 | 23.86 | 3.00 | 4.331 | 1.499 |
| 6 | 2247 | 100 | 2147 | 0.0445 | 100 | 89 | 0 | 771 | 0 | 1287 | 0.53 | 1.00 | 23.86 | 5.99 | 4.331 | 2.445 |
| 7 | 3147 | 1000 | 2147 | 0.31766 | 1000 | 89 | 0 | 771 | 0 | 1287 | 0.92 | 1.00 | 23.87 | 59.94 | 4.332 | 5.589 |
| 8 | 4294 | 2147 | 2147 | 0.5 | 2147 | 89 | 0 | 771 | 0 | 1287 | 0.96 | 1.00 | 23.88 | 599.16 | 4.332 | 8.733 |

#### Recommendation 5: Evidence towards benignity can be allocated where there are sufficient concordant pathogenic missense truthset variants; alternatively where there is signal of disease association for assay-defined deleterious variants en-masse, a single EP may be allocated

- Pathogenic truthset variants should be of penetrance typical for that gene and act via the mechanism established for the relevant disease phenotype (as much as available information permits).
- Evidence towards benignity (1EP) can be allocatable where there are three or more fully concordant pathogenic missense truthset variants as per the ClinGen framework.
- Where fewer than three pathogenic missense truthset variants exist, one EP for benignity may be allocatable on the basis of ‘en masse association’ with disease of assay-defined deleterious (non-functional) missense variants where:

`o` the gene is understood to have a single mechanism of pathogenicity, which is measured by the assay under evaluation,
`o` all known pathogenic truthset missense variants have concordant deleterious (non-functional) assay readouts,
`o` PTVs have concordant deleterious (non-functional) assay readouts, and
`o` association is demonstrated between assay-defined deleterious (non-functional) missense variants as a group and the relevant disease phenotype. A statistically significant (p<0.05) odds ratio (OR) of clinically-appropriate magnitude must be demonstrated.

#### Recommendation 6: The distribution of assay scores for missense truthset variants should be examined

Where the assayist-defined thresholds for pathogenic and benign evidence utilised PTV/synonymous variant sets, it is informative to compare the distribution and median assay scores for PTV/synonymous versus missense truthsets. Where the distributions differ, caution is required regarding the thresholds on account of (i) greater potential for misclassification of variants near the threshold, along with (ii) attenuation of EPs for pathogenicity or benignity (see Box 2). High stringency missense truthset variants are most informative for this comparison.

#### Recommendation 7: To reduce circularity regarding truthsets classified with functional data, appropriate filtering or censoring of ClinVar should be attempted

Aspirational best practice would dictate (i) all truthset variants should have been attained clinical classification as (likely) pathogenic/benign without the contribution of functional data with (ii) censoring of ClinVar at a date prior to the assay publication to avoid direct circularity involving the assay under evaluation. However, time-censoring applied as routine could be unduly punitive in disqualifying all recent classifications, regardless of inclusion of the assay data under evaluation.

###### Box 2: Demonstration of threshold calibration using PTV/synonymous variants, where there is a scoring zone absent of variant data

The thresholds (shown here with red and blue lines, with the area in between thus representing the ‘intermediate’ (grey) zone) is typically assigned according to a maximum-likelihood estimation to separate all PTV/deleterious variants from all synonymous/neutral variants. Where there is a zone ‘bald’ of either variant type, theoretically the ‘true’ threshold for missense calibration could lie at any point between the highest scoring PTV and the lowest scoring synonymous variant. We illustrate for an assay how score distributions for pathogenic and benign missense truthsets might differ from those for PTV/synonymous variants, and how for such underlying distributions clinical validation using missense truthsets applying these thresholds will only serve to under-score the allocatable EPs for pathogenicity/benignity.

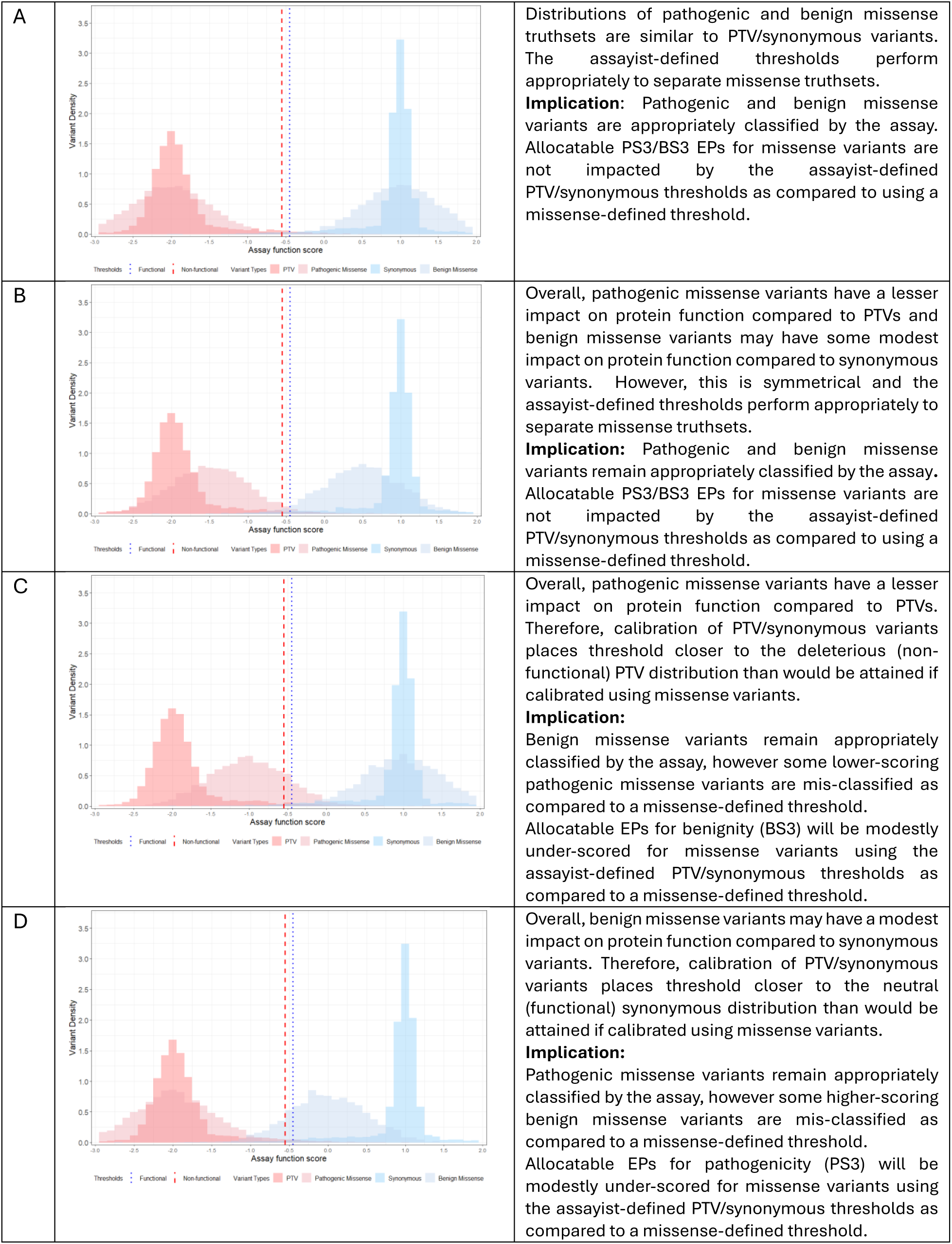

The following ancillary recommendations are applicable for specific scenarios, and should be considered in the context of assay under review:

#### Ancillary recommendation 1: Where the assay does not recapitulate genomic context (eg cDNA-based assays)

For cDNA-based assays, there should be caution in ascribing EPs for benignity for variants where deleterious impact on protein function will not typically be captured by the assay, namely nonsense, potentially spliceogenic and noncoding variants. Accordingly, for cDNA-based assays, pathogenicity truthsets should only include missense variants with a non-splicing mode of action.

#### Ancillary recommendation 2: Where the assay reports multiple deleterious ‘zones’

Clinical validation may proceed using one of the following methodologies:

- Combination of zones (for example, combining all deleterious zones into one zone)
- Using the most extreme scoring zone(s) only and treating less extreme zones as intermediate
- Evaluation of each zone separately using an independent phenotype-specific pathogenic truthset, where zones are suspected to be associated with distinct phenotypes. Gene-specific expertise will be critical for appraising such scenarios.

Whilst scoring zones may be described as indicating intermediate or reduced penetrance, truthsets of variants of clinically-verified intermediate/reduced penetrance do not currently exist, meaning validation against this variant status is not currently possible.

#### Ancillary recommendation 3: Where the assay reports multiple readouts

Where multiple readouts are reported for each variant, it will be necessary to determine upfront whether (i) the readouts should be harmonised into a composite score upstream of clinical validation or (ii) these outputs should be treated in manner of being standalone assays. Gene-specific expertise will be critical for appraising these scenarios, but as broad guidance:

- **Scenario 1: the multiple readouts are technical duplicates.** These values should, following quality control checks, be averaged to provide a consensus measurement (this is typically performed by the assayist).
- **Scenario 2: the multiple readouts are from multiple assays measuring an identical function**, for example for HRR genes assays measuring sensitivity to multiple different PARP inhibitors(22). In this situation, multiple readouts could be harmonised upstream into a single assignation by requiring concordance ie deleterious/deleterious or neutral/neutral whereby otherwise the readout is assigned as indeterminate
- **Scenario 3: multiple readouts are from different methods for measurement of the same function,** for example cell survival and protein binding as readouts of DNA repair. In this situation, multiple readouts could be harmonised upstream into a single assignation with a variant being ascribed as deleterious (non-functional) if either readout is deleterious, but for benignity requiring both readouts to be neutral (functional).
- **Scenario 4: multiple readouts are from measurements of different mechanisms of pathogenicity,** for example assays of TP53 in which both gain of function and loss of function are quantified. In this scenario where different mechanisms are recognised a priori, aspirational best practice would be that separate validations should be undertaken using different truthsets reflecting the different mechanisms of action. Operationally, this will be highly challenging to unpick outside of genes already well-characterised a priori as having different mechanisms of action.

## Discussion

Aligning with the ClinGen assay-level clinical validation framework from Brnich et al and recognising the predominant use-case for functional data being resolution of rare missense VUS, there was clear consensus within CanVIG-UK that missense variants should be used for clinical validation in assays of cancer susceptibility genes.

However, within CanVIG-UK we also recognised the challenges inherent to this approach: namely a frequent paucity of both benign and (for many genes) pathogenic high-stringency ClinVar-classified missense variants. Our extensive analyses of different truthsets provided an evidence basis by which we advocate for relaxation of stringency for truthset construction. This includes relaxing the number of ClinVar stars, permitting of classifications with soft-conflicts and inclusion of “likely” pathogenic and “likely” benign classifications.

Where, despite this, there remains a paucity of benign missense variants, this limits the allocation of assay evidence towards pathogenicity, the most important clinical priority (Principle C). We propose this paucity can be mitigated by construction and integration of proxy-clinical benign-classified missense variants to bolster truthset size. These are variants which have not been reported in ClinVar as identified in a clinical setting, but instead are systematically classified following the standards set by the ACMG-AMP with automatic application of evidence to reach a classification of (likely) benign, via leveraging of a gene-specific maximum tolerated allele frequency (MTAF) in combination with in silico predictions using gene-specific thresholds (11)(23). In the context of very rare highly-penetrant early-onset disease, the MTAF principle might be extended such that any missense variant observed in a population database such as gnomAD(24) might be ascribed as a proxy-clinical benign missense truthset variant.

For many genes, paucity of pathogenic missense variants is less problematic, especially if stringency is relaxed. Furthermore, (i) strengthening of a variant’s evidence towards benignity tends to be less of a clinical priority and (ii) fewer EPs are required. Nevertheless, it is important to demonstrate that the assay can correctly identify pathogenic missense variants as deleterious (non-functional): this is central to being confident in assay allocation of evidence for benignity (BS3). At least three concordant ClinVar-classified pathogenic truthset variants are required to attain one EP for benignity using the ClinGen framework. Should there be an extreme paucity of ClinVar-classified pathogenic variants, we concurred that a single evidence point towards benignity could be awarded where the assay-defined deleterious variants *en masse* exhibit significant association with disease.

Within the upcoming ACMG/AMP/CAPP/ClinGen v4.0 framework, clinical validation using a quantitative evidence strength scale will potentially enable application of evidence up to +8/-8, and classification of likely pathogenic or benign can be attained with functional data alone. Indeed, for many of the gene-assay combinations we have explored, the highest EP for pathogenicity exceeded the +6 points required for a Likely Pathogenic classification if PTV/synonymous or ‘All ClinVar variant’ truthsets were used (Figure 1). Furthermore, with the increase in unaffected/population testing, attention is urgently required regarding use of functional data as the sole evidence for classification where the variant is observed in an unaffected individual in the absence of historic clinical observations.

### Limitations and Priorities for future CanVIG-UK work

In development of these recommendations within CanVIG-UK, we identified the following as limitations of the current approaches to clinical validation of functional assays and priorities for additional work and guidance:

#### Calibration-validation models affording graded evidence allocation

We recognised the inherent limitations of the ClinGen assay-level clinical validation framework described by Brnich et al insofar as it generates a single evidence score for pathogenicity and a single evidence score for benignity. There is reliance on the assayist-defined thresholds, derivation of which may not be transparent and may have used PTVs and synonymous variants, the score distribution for which may vary compared to that of missense variants. Due to its dichotomous output of EPs, this assay-level clinical validation approach may serve to (i) under-allocate EPs for high-scoring variants and (ii) over-allocate EPs for variants scoring very near the thresholds.

These issues can in principle be addressed by undertaking simultaneous de novo calibration and validation, modelling a range of variant scores and concomitant thresholds (18, 25, 26). However, such approaches also have challenges. Firstly, the truthset size issue may be even more problematic as these models are highly unstable with small truthset numbers especially over the intermediate scoring zones; as with any model, best practice would involve training and testing using separate truthsets. Secondly, such models are inherently less transparent than the current assay-level approach and require more specialist technical resource, precluding ready reproduction by different groups of diagnostic scientists: routes to clinical adoption will thus be commensurately more challenging (13).

#### Proxy-clinical pathogenic-classified missense variants

While we have developed methodology and guidance for construction of ‘proxy-clinical’ benign-classified missense variant truthsets(11), there is no equivalent approach for generating pathogenicity truthset variants. Whilst functional data suggest they are deleterious to protein impact, for a number of cancer susceptibility genes there is a dramatic lack of clinically-classified pathogenic missense variants, for example *PALB2* and *BARD1*. Whilst we have proposed ‘en-masse-association’ of assay-assigned deleterious missense variants with disease can be used for judicious allocation of a single benignity EP, more sophisticated quantitative approaches are required for proxy-clinical pathogenic-classified missense truthsets.

#### Assay combinations

Although we have delineated conceptual scenarios whereby multiple readouts may exist for the same variant, defining which scenario is pertinent may in practice be challenging. With the exponential increase of MAVE data availability, robust methodology is urgently required for (i) robust delineation of whether assays are measuring different functions or whether they are discordant due to differential assay quality, and (ii) appropriate allocation of evidence where the assay readouts concur and, separately, where they conflict(27). Having sufficient appropriate truthset variants to generate testing *and* training sets would provide most flexibility, but as previously, availability of truthset variants will be operationally limiting for all but a minority of genes.

#### Reduced penetrance variants

It has been posed that variants with a reduced deleterious score may be of intermediate or reduced penetrance(20). As yet, these findings cannot be clinically validated as consensus clinically-classified truthsets of reduced penetrance variants do not exist, for even the best-studied genes. However, work cross-iterating validations of case-control, segregation, splicing, in silico and functional data may provide improved routes to demonstration of reduced penetrance with concomitant classifications and accordant availability of truthsets.

#### Non-coding (spliceogenic) variants

As assays are now interrogating variants deeper into introns, additional work will be required to evaluate generalisability to these variant types of current clinical validation based on missense variants.

## Conclusion

Previous work has demonstrated an implementation gap between publication of new functional data and clinical implementation of assay data in the UK and more widely. Via the CanVIG-UK network and overarching CStAG group, we have (i) articulated principles underpinning our approach to clinical validation practice and truthset construction, (ii) agreed best practice recommendations for truthset construction which build on the ClinGen assay-level clinical validation framework, and (iii) established priorities for future work around clinical validation of functional assays to further address the implementation gap.

## Data Availability

All data generated or analysed during this study are included in this published article [and its supplementary information files].

## Declarations

### Ethics approval and consent to participate

Not applicable

### Consent for publication

Not applicable

### Authors’ contributions

Conceptualisation: CT, AS, FPR, LS, DA, GF; Data Curation: SA; Software: CFR, SA; Formal Analysis: SA, CFR; Funding acquisition: CT; Investigation: SA; Methodology: CT, SA, CFR, AG; Project Administration: SA, Supervision: CT; Visualisation: SA; Validation: MD, GJB, AC, RR, JF, BF, SPS, JG, JP, EJ, TMcD, LH, LY-S, PL, LR, KS, HH, TMcV, CanVIG-UK; Writing – original draft: CT, SA; Writing - review and editing: all authors.

## References

1. McEwen AE, Tejura M, Fayer S, Starita LM, Fowler DM. Multiplexed assays of variant effect for clinical variant interpretation. Nature reviews Genetics. 2026;27(2):137–54.

2. Fowler DM, Rehm HL. Will variants of uncertain significance still exist in 2030? American journal of human genetics. 2024;111(1):5–10.

3. Dawood M, Fayer S, Pendyala S, Post M, Kalra D, Patterson K, et al. Using multiplexed functional data to reduce variant classification inequities in underrepresented populations. Genome medicine. 2024;16(1):143.

4. Richards S, Aziz N, Bale S, Bick D, Das S, Gastier-Foster J, et al. Standards and guidelines for the interpretation of sequence variants: a joint consensus recommendation of the American College of Medical Genetics and Genomics and the Association for Molecular Pathology. Genetics in medicine : official journal of the American College of Medical Genetics. 2015;17(5):405–24.

5. Ellard S, Baple EL, Owens M, Cannon S, Turnbull C, Eccles D, et al. ACGS Best Practice Guidelines for Variant Classification 2018. Association for Clinical Genetics Science (ACGS); 2018.

6. Garrett A, Callaway A, Durkie M, Cubuk C, Alikian M, Burghel GJ, et al. Cancer Variant Interpretation Group UK (CanVIG-UK): an exemplar national subspecialty multidisciplinary network. Journal of medical genetics. 2020.

7. Garrett A, Allen S, Rowlands CF, Choi S, Durkie M, Burghel GJ, et al. Cancer Variant Interpretation Group UK (CanVIG-UK): updates on an exemplar national subspecialty multidisciplinary network. medRxiv. 2026.

8. Tavtigian SV, Greenblatt MS, Harrison SM, Nussbaum RL, Prabhu SA, Boucher KM, et al. Modeling the ACMG/AMP variant classification guidelines as a Bayesian classification framework. Genetics in medicine : official journal of the American College of Medical Genetics. 2018;20(9):1054–60.

9. Tavtigian SV, Harrison SM, Boucher KM, Biesecker LG. Fitting a naturally scaled point system to the ACMG/AMP variant classification guidelines. Human mutation. 2020.

10. Brnich SE, Abou Tayoun AN, Couch FJ, Cutting GR, Greenblatt MS, Heinen CD, et al. Recommendations for application of the functional evidence PS3/BS3 criterion using the ACMG/AMP sequence variant interpretation framework. Genome medicine. 2019;12(1):3.

11. Rowlands CF, Allen S, Garrett A, Durkie M, Burghel GJ, Robinson R, et al. Availability of benign missense variant “truthsets” for validation of functional assays: current status and a novel systematic approach. 2025.

12. Allen S, Garrett A, Muffley L, Fayer S, Foreman J, Adams DJ, et al. Workshop report: the clinical application of data from multiplex assays of variant effect (MAVEs), 12 July 2023. European journal of human genetics : EJHG. 2024;32(5):593–600.

13. Allen S, Garrett A, Rowlands CF, Durkie M, Burghel GJ, Robinson R, et al. Validating data from multiplex assays of variant effect: A CanVIG-UK national survey of NHS clinical scientists. American journal of human genetics. 2025;112(6):1479–88.

14. Villani RM, Terrill B, Tudini E, McKenzie ME, Cliffe CC, Hahn CN, et al. Consultation informs strategies for improving the use of functional evidence in variant classification. American journal of human genetics. 2025;112(6):1489–95.

15. Park MS, Kumar RD, Ovadiuc C, Folta A, McEwen AE, Snyder A, et al. Insights on improving accessibility and usability of functional data to unlock their potential for variant interpretation. American journal of human genetics. 2025;112(6):1468–78.

16. Allen S, Rowlands CF, Kuzbari Z, Garrett A, Durkie M, Burghel GJ, et al. Clinical validation of large-scale functional assays: insights from 2,120 gene-truthset-assay evaluations. medRxiv, Manuscript in Draft. 2026.

17. Findlay GM, Daza RM, Martin B, Zhang MD, Leith AP, Gasperini M, et al. Accurate classification of BRCA1 variants with saturation genome editing https://sge.gs.washington.edu/BRCA1/. Nature. 2018;562(7726):217–22.

18. Huang H, Hu C, Na J, Hart SN, Gnanaolivu RD, Abozaid M, et al. Functional evaluation and clinical classification of BRCA2 variants. Nature. 2025;638(8050):528–37.

19. Sahu S, Galloux M, Southon E, Caylor D, Sullivan T, Arnaudi M, et al. Saturation genome editing-based clinical classification of BRCA2 variants. Nature. 2025;638(8050):538–45.

20. Olvera-León R, Zhang F, Offord V, Zhao Y, Tan HK, Gupta P, et al. High-resolution functional mapping of RAD51C by saturation genome editing. Cell. 2024;187(20):5719–34.e19.

21. Buckley M, Terwagne C, Ganner A, Cubitt L, Brewer R, Kim DK, et al. Saturation genome editing maps the functional spectrum of pathogenic VHL alleles. Nature genetics. 2024;56(7):1446–55.

22. Bouwman P, van der Heijden I, van der Gulden H, de Bruijn R, Braspenning ME, Moghadasi S, et al. Functional Categorization of BRCA1 Variants of Uncertain Clinical Significance in Homologous Recombination Repair Complementation Assays. Clinical cancer research : an official journal of the American Association for Cancer Research. 2020;26(17):4559–68.

23. Whiffin N, Minikel E, Walsh R, O’Donnell-Luria AH, Karczewski K, Ing AY, et al. Using high-resolution variant frequencies to empower clinical genome interpretation. Genetics in medicine: official journal of the American College of Medical Genetics. 2017.

24. Karczewski KJ, Francioli LC, Tiao G, Cummings BB, Alföldi J, Wang Q, et al. The mutational constraint spectrum quantified from variation in 141,456 humans. Nature. 2020;581(7809):434–43.

25. Badonyi M, Marsh JA. acmgscaler: an R package and Colab for standardized gene-level variant effect score calibration within the ACMG/AMP framework. Bioinformatics (Oxford, England). 2025;41(10).

26. Zeiberg D, Tejura M, McEwen AE, Fayer S, Pejaver V, Rubin AF, et al. Gene-based calibration of high-throughput functional assays for clinical variant classification. bioRxiv. 2025.

27. Calhoun JD, Dawood M, Rowlands CF, Fayer S, Radford EJ, McEwen AE, et al. Combining multiplexed functional data to improve variant classification. ArXiv. 2025.

