## Supplementary Table 1 for "‘Truthsets’ for clinical validation of large-scale functional assays: Practice recommendations from Cancer Variant Interpretation Group UK (CanVIG-UK)"

### Supplementary Table 1: Truthset source for the highest scoring missense assay in each gene-assay combination presented in Figure 1

| **Gene/Assay** | **Score** | **Code** | **Variant Type** | **Source (pathogenic)** | **Source (benign)** | **ClinVar class** | **ClinVar Soft Conflicts** | **ClinVar stars** | **No. pathogenic controls** | **No. benign controls** |
| --- | --- | --- | --- | --- | --- | --- | --- | --- | --- | --- |
| BRCA1 | 4.94 | EP_PS3 | Missense + Proxy-clinical benign | ClinVar | ClinVar + Proxy-clinical set 4 | P/LP and B/LB | No | ≥2 | 44 | 1107 |
| BRCA1 | -4.23 | EP_BS3 | Missense | ClinVar | ClinVar | P/LP and B/LB | Yes | Any | 91 | 40 |
| BRCA1 | -4.23^a^ | EP_BS3 | Missense + Proxy-clinical benign | ClinVar | ClinVar + Proxy-clinical set 1 | P/LP and B/LB | Yes | Any | 91 | 40 |
| BRCA2 (Huang et al) | 3.88 | EP_PS3 | Missense | ClinVar | ClinVar | P/LP and B/LB | Yes | ≥1 | 116 | 310 |
| BRCA2 (Huang et al) | 3.88 | EP_PS3 | Missense | ClinVar | ClinVar + Proxy-clinical set 1 | P/LP and B/LB | Yes | ≥1 | 116 | 310 |
| BRCA2 (Huang et al) | -4.21 | EP_BS3 | Missense + Proxy-clinical benign | ClinVar | ClinVar | P/LP and B/LB | Yes | ≥1 | 116 | 310 |
| BRCA2 (Huang et al) | -4.21 | EP_BS3 | Missense + Proxy-clinical benign | ClinVar | ClinVar + Proxy-clinical set 1 | P/LP and B/LB | Yes | ≥1 | 116 | 310 |
| BRCA2 (Sahu et al) | 3.86 | EP_PS3 | Missense + Proxy-clinical benign | ClinVar | ClinVar + Proxy-clinical set 1 | P/LP and B/LB | No | Any | 70 | 58 |
| BRCA2 (Sahu et al) | -3.11 | EP_BS3 | Missense | ClinVar | ClinVar | P/LP and B/LB | Yes | ≥1 | 121 | 321 |
| BRCA2 (Sahu et al) | -3.11^a^ | EP_BS3 | Missense + Proxy-clinical benign | ClinVar | ClinVar + Proxy-clinical set 1 | P/LP and B/LB | Yes | ≥1 | 121 | 321 |
| RAD51C | 3.56 | EP_PS3 | Missense + Proxy-clinical benign | ClinVar | ClinVar + Proxy-clinical set 2 | P/LP and B/LB | Yes | Any | 29 | 126 |
| RAD51C | 3.56 | EP_PS3 | Missense + Proxy-clinical benign | ClinVar | ClinVar + Proxy-clinical set 2 | P/LP and B/LB | Yes | ≥1 | 29 | 126 |
| RAD51C | -2.93 | EP_BS3 | Missense + Proxy-clinical benign | ClinVar | ClinVar + Proxy-clinical set 1 | P/LP and B/LB | No | Any | 9 | 20 |
| RAD51C | -2.93^a^ | EP_BS3 | Missense + Proxy-clinical benign | ClinVar | ClinVar + Proxy-clinical set 1 | P/LP and B/LB | No | ≥1 | 9 | 20 |
| VHL | 4.30 | EP_PS3 | Missense + Proxy-clinical benign | ClinVar | ClinVar + Proxy-clinical set 4 | P/LP and B/LB | No | ≥2 | 83 | 206 |
| VHL | -3.99 | EP_BS3 | Missense + Proxy-clinical benign | ClinVar | ClinVar + Proxy-clinical set 4 | P/LP and B/LB | No | ≥2 | 83 | 206 |

EP_PS3 = highest scoring truthset for evidence points in favour of pathogenicity. EP_BS3 = highest scoring truthset for evidence points in favour of benignity. Truthsets containing only missense variants are included in this table, following the best practices described in this paper. Where there are multiple missense truthset options which provide the highest score for an assay, both are listed. Proxy-clinical sets were defined as follows: Set 1 = Attains BA1 in any non-founder population in gnomAD v4.1; Set 2 = Attains BS1_strong in any non-founder population in gnomAD v4.1 and benign evidence from either REVEL (≤0.290) or BayesDel noAF (≤0.15 for BRCA1 and ≤0.18 for BRCA2, ≤-0.18 for VHL and RAD51C); Set 3 = Attains BS1_sup in any non-founder population in gnomAD v4.1 and benign evidence from at least one predictive tool (did not produce highest scoring gene-assay combination for any assay examined); Set 4 = Attains BS1_sup in any non-founder population in gnomAD v4.1 and/or benign evidence from at least one predictive tool. A ‘soft conflict’ is defined as a variant classified in ClinVar as a VUS and as either (likely) pathogenic or (likely) benign.

^a^Addition of proxy-clinical benign variants is anticipated to impact EPs towards pathogenicity (EP_PS3), but have a negligible impact on EPs towards benignity (EP_BS3). The exception is where there were no concordant benign variants from the ClinVar-derived truthsets, and addition of the proxy-clinical set enabled EP_BS3 calculation per the ClinGen assay-level clinical validation framework based on the available pathogenic missense truthset variants (see Recommendation 4).
